# Retention in care and viral suppression after same-day ART initiation: One-year outcomes of the SLATE I and II individually randomized clinical trials in South Africa

**DOI:** 10.1101/2021.06.11.21258784

**Authors:** Mhairi Maskew, Alana T Brennan, Matthew P Fox, Lungisile Vezi, Willem DF Venter, Sydney Rosen

## Abstract

**Introduction:** Same-day initiation (SDI) of antiretroviral therapy (ART) for HIV consistently increases ART uptake, but concerns remain about higher attrition from care after initiation. We analysed twelve-month retention in the SLATE SDI trials.

**Methods:** SLATE I and SLATE II were individually randomized trials at public outpatient clinics in Johannesburg that enrolled patients not yet on ART and administered the SLATE I or II algorithm, which included a symptom self-report, medical history, brief physical examination, and readiness questionnaire, to assess eligibility for SDI. ART uptake and early retention have been reported. Using routine clinic records, we conducted a pooled analysis of retention in care and HIV viral suppression 14 months after study enrolment.

**Results and discussion:** We enrolled 1,193 study participants (standard arms, n=599, 50%; intervention arms, n=594, 50%) and analysed by originally assigned groups. By 14 months after study enrolment (equivalent to 12-month retention in care), 50% of intervention arm patients and 46% of standard arm patients remained in care at the initiating site (crude risk difference 4% (95% confidence interval −1% to 10%; crude relative risk 1.10 (0.97-1.23), with similar viral suppression between arms. Observed attrition from care at site by 14 months was high in both study arms, but we found no evidence that the offer of SDI led to greater overall attrition or lower rates of viral suppression one year after starting ART and may have led to small improvements. Same-day initiation may have shifted some attrition from before to after dispensing of the first dose of medication.

**Conclusions:** An offer of same-day initiation of ART, following a carefully designed protocol to identify patients who are eligible and ready to start treatment, is not inherently associated with an overall increase in patient attrition from care.

**Trial registration:** Clinicaltrials.gov NCT02891135, registered September 1, 2016. First participant enrolled March 6, 2017 in South Africa. Clinicaltrials.gov NCT03315013, registered October 19, 2017. First participant enrolled 14 March 2018.

## INTRODUCTION

Since 2017, when the World Health Organization recommended rapid initiation of antiretroviral therapy (ART) for people living with HIV and “same-day initiation” of ART for patients who are ready for treatment on the day they test positive for HIV [1], many countries in sub-Saharan Africa, including South Africa [2], have introduced the possibility of same-day ART initiation into their national HIV programs. To help guide decisions on exactly who should be eligible for same-day initiation (SDI) and how to implement it, we developed and evaluated two algorithms in South Africa. SLATE I (Simplified Algorithms for Treatment Eligibility I) [3] and SLATE II [4] were designed as simple, clinical algorithms that require no point-of-care laboratory testing and can be used by existing healthcare personnel to distinguish patients who can start ART that day, even if they have mild symptoms of illness, from those who do require additional care prior to initiation.

While evidence from the SLATE trials [5,6] and others [7,8] demonstrate improved uptake of ART with SDI compared to standard care, concerns remain whether the benefits of SDI can be translated into improved retention once on treatment, or if instead attrition is simply shifted from before to soon after ART initiation, or even made worse by pressure that the expectation of same-day initiation is perceived to place on patients [9,10]. Overall attrition at 8 months was lower in the intervention arm in both trials [5,6], but its timing differed. In SLATE I, roughly one third of the attrition observed in the standard arm but more than half in the intervention arm occurred after patients initiated ART; in SLATE II, half of the attrition observed in the standard arm but nearly three quarters of the attrition observed in the intervention arm occurred after initiation. A limitation of both studies was that the primary outcome—a combined indicator of initiation of ART within 28 days and retention in care 8 months after study enrolment—was assessed at a time representing just six months on ART. Full 12-month retention outcomes remain unclear. We present SLATE I and SLATE II retention and viral suppression outcomes 14 months after study enrolment (approximately 12 months after ART initiation) to determine whether the differences between arms observed at 8 months persisted to 14 months.

## METHODS

### Study design

SLATE I and SLATE II were individually randomised, non-blinded pragmatic evaluations to assess the effect of each SLATE algorithm on ART initiation and retention in care. Both studies have been described in detail elsewhere [3–6,11,12]. Both algorithms consisted of four screening tools (Figure 1), each evaluating specific criteria for same-day ART initiation: 1) symptom report, 2) medical history, 3) physical examination, and 4) patient readiness assessment. Intervention arm patients found to be eligible on all four screening tools were offered initiation of ART on the day of study enrolment. Those ineligible on any of the screens were referred back to routine care at the study site clinic for further services prior to ART initiation; clinics could still offer ART initiation that same day if they chose.

**Figure 1.**
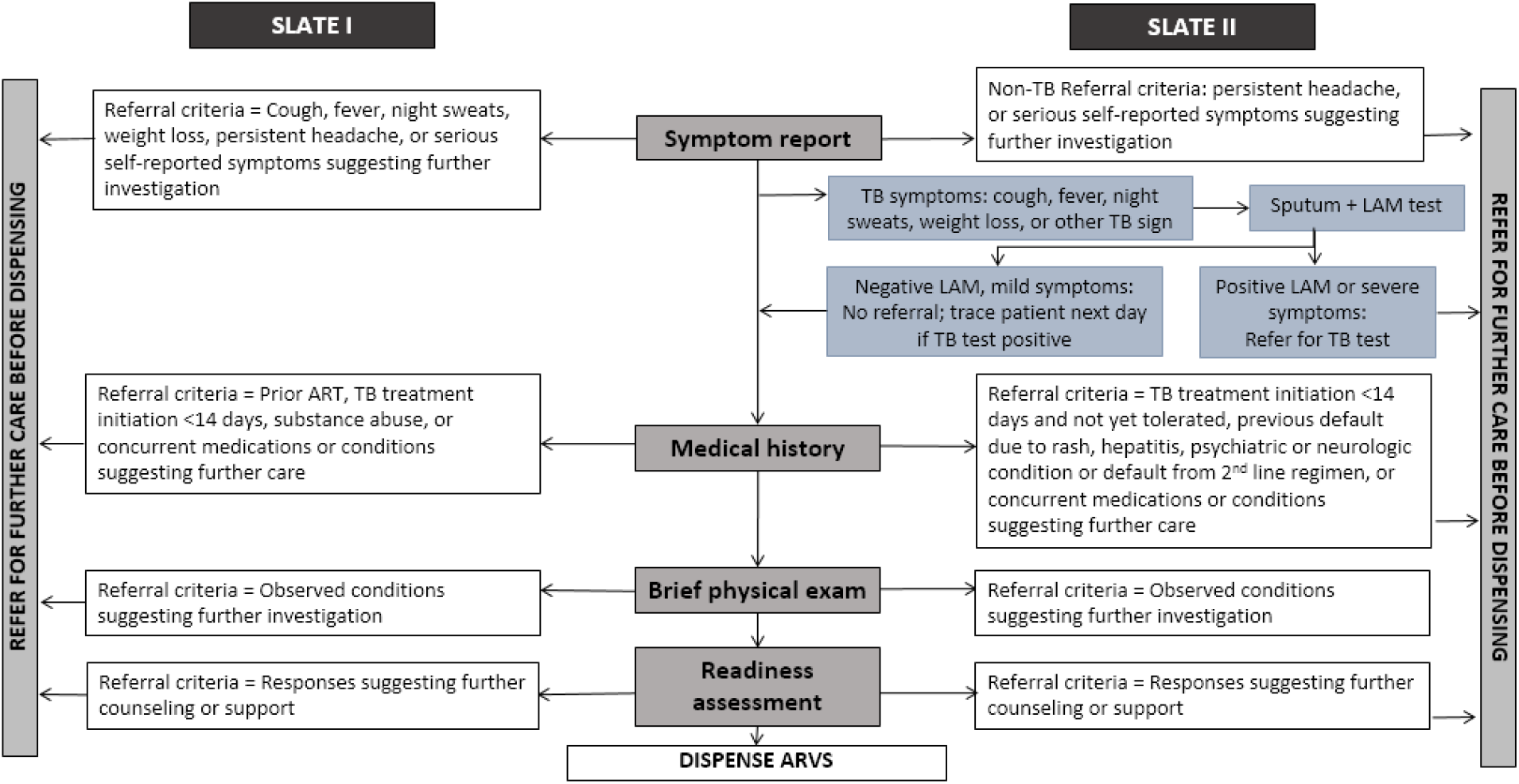
SLATE I and SLATE II algorithms.

### Study setting and population

SLATE I was conducted in Kenya and South Africa and SLATE II in South Africa. We report data from both studies for the South Africa sites only, as the Kenya sites did not continue follow up through 14 months. The study sites were high-volume, public-sector primary care clinics in urban formal and informal settings around Johannesburg, South Africa. Care was provided according to the relevant South African National Department of Health ART guidelines in place during each study [13]. Non-pregnant, HIV infected adults ≥ 18 years presenting at the study sites for HIV diagnosis or any type of HIV care but not yet on ART were eligible for the studies.

### Study procedures

Consented and enrolled study participants completed an interviewer-administered questionnaire and were randomized 1:1 to either the intervention arm (SLATE algorithm) or standard arm (routine clinic procedures). After randomization, standard arm patients continued their clinic visits under standard of care. Patients randomized to the intervention arms were referred to a study nurse who administered the four SLATE algorithm screens and then dispensed ART directly to those patients eligible for SDI and referred back to the facility those patients requiring additional services prior to initiation of ART. Patients in both arms were followed passively, through medical record review, and had no further interaction with study staff.

### Study outcomes and data analysis

We report here the secondary outcomes of 1) initiation of ART within 28 days and retention on ART 14 months after study enrolment and 2) suppression of viral load (to <400 copies/mL) by 14 months after study enrolment. A patient was considered retained if we observed a clinic visit or laboratory test in the patient’s medical record between 11 and 14 months after study enrolment. Fourteen months was selected to allow up to 1 month to initiate ART, 12 months of follow up after treatment initiation, and up to 1 month to return for the 12-month routine clinic visit. Patients with no evidence of a clinic visit or laboratory test during this period were assumed lost to follow up.

All follow-up data were sourced from routinely collected medical records in TIER.Net [14], South Africa’s national HIV monitoring system, and supplemented with routine electronic and paper records at the site and laboratory records from the National Health Laboratory System (NHLS). For this analysis, we pooled both study samples and conducted a crude analysis comparing the proportion of patients achieving each dichotomous outcome by study arm. We estimated crude risk ratios and crude risk differences and their corresponding 95% confidence intervals (CI) for each outcome, by intention-to-treat.

## Ethics

The studies were approved by the Human Research Ethics Committee of the University of the Witwatersrand (Medical) and the institutional review board of Boston University Medical Campus.

## RESULTS AND DISCUSSION

### Summary of prior findings

Participants were enrolled between March 6 and July 28, 2017 for SLATE I and between 14 March and 18 September 2018 for SLATE II. As previously reported, a total of 1,193 study participants in both studies were randomized to the standard arm (n=599, 50%) or the intervention arm (n=594, 50%) and analysed by originally assigned groups. The study population was predominantly female (63%) with a median age of 35 years (IQR 29-41) and median CD4 count of 293 cells/mm^3^ (133-487). No important imbalances with respect to the characteristics of the enrolled sample were noted.

In the combined sample of 1,193, a total of 447/599 standard arm patients (74.6%) and 509/594 intervention arm patients (85.7%) initiated ART within 28 days of study enrolment. Achievement of the combined primary outcome of initiation within 28 days and retention in care at 8 months was 321/599 (53.6%) and 381/594 (64.1%) for the standard and intervention arms, respectively, revealing very high rates of attrition from these sites—46.4% and 35.9%—by 8 months after study enrolment.

### Retention and suppression at 14-month endpoint

In Table 1, we report outcomes at 14 months after study enrolment. Initiation within 28 days and retention at 8 months are included in Table 1 for comparison. Separate results for each study are provided in Supplemental Tables 1 and 2.

**Table 1:** Outcomes at 14 months after study enrolment by arm (n= 1193)

| Outcome | Standard arms<br>(n=599) | Intervention<br>arms (n=594) | Crude RD<br>(95%CI) † | Crude RR (95% CI) † |
| --- | --- | --- | --- | --- |
| **Previously reported outcomes** |  |  |  |  |
| Initiated ART ≤ 28 days of study enrolment | 447 (75%) | 509 (86%) | 11% (7 to 16%) | 1.15 (1.08-1.22) |
| Initiated ART ≤ 28 days <u>and</u> retained in care<br>8 months after study enrolment | 321 (54%) | 381 (64%) | 10% (5 to 16%) | 1.20 (1.09-1.32) |
| Initiated ART ≤ 28 days <u>and</u> known to be<br>virally suppressed by 8 months | 184 (31%) | 223 (38%) | 7% (1 to 12%) | 1.22 (1.04-1.43) |
| **14-month outcomes (retention)\*** |  |  |  |  |
| Initiated ART ≤ 28 days <u>and</u> retained in care<br>14 months after study enrolment | 275 (46%) | 299 (50%) | 4% (-1 to 10%) | 1.10 (0.97-1.23) |
| Initiated ART ≤ 28 days, not retained 14<br>months after study enrolment | 173 (29%) | 210 (35%) | 6% (1 to 12%) | 1.22 (1.04-1.45) |
| Did not initiate ≤ 28 days | 151 (25%) | 85 (14%) | -11% (-15 to 6%) | 0.57 (0.45-0.72) |
| **14-month outcomes (viral suppression)\*\*** |  |  |  |  |
| Initiated ART ≤ 28 days <u>and</u> known to be<br>virally suppressed by 14 months | 163 (27%) | 173 (29%) | 2% (-3 to 7%) | 1.07 (0.89-1.28) |
| Initiated ART ≤ 28 days <u>and</u> known to be<br>virally unsuppressed by 14 months | 15 (3%) | 14 (2%) | 0% (-2 to 2%) | 0.94 (0.46-1.93) |
| No viral load test results found | 97 (16%) | 112 (19%) | 3% (-2 to 8%) | 1.16 (0.91-1.49) |
| **Alternate 14-month outcomes (viral**<br>**suppression)\*\*\*** |  |  |  |  |
| Initiated ART ≤ 28 days <u>and</u> known to be<br>virally suppressed by 14 months | 181 (30%) | 199 (34%) | 4% (-2% to 9%) | 1.11 (0.94-1.31) |
| Initiated ART ≤ 28 days <u>and</u> known to be<br>virally unsuppressed by 14 months | 18 (3%) | 18 (3%) | 0% (-2 – 2%) | 1.00 (0.53-1.91) |
| No viral load test results found | 76 (13%) | 82 (13%) | 1% (-3% - 5%) | 1.09 (0.81-1.46) |
| **Alternate viral load outcomes<sup>‡</sup>** |  |  |  |  |
| Any suppressed viral load after study<br>enrolment | 297 (50%) | 320 (54%) | 4% (-1% to 10%) | 1.09 (0.97-1.21) |
| Any unsuppressed viral load result after<br>study enrolment | 37 (6%) | 38 (6%) | 0% (-3% to 3%) | 1.04 (0.67-1.61) |
| No viral load results observed | 265 (44%) | 236 (40%) | -4% (-10% to 1%) | 0.90 (0.79-1.03) |

†Reference group: standard arm
\*Per protocol outcome definition: Observed clinic visit or VL test between months 11-14 after study enrolment
\*\*Per protocol outcome definition: Observed VL test between months 11-14 after study enrolment
\*\*\*Alternate outcome definition: Observed VL test between months 9-14 after study enrolment
<sup>‡</sup>Alternate outcome definition: Any viral load test result observed at any point up to 14 months after study enrolment regardless of whether the patient initiated ART or not

As Table 1 indicates, retention in care at the study sites by 14 months was poor in both study arms. The numerical advantage seen in the intervention arms of the trials at the 8-month primary endpoints, however, persisted through 14 months, when 50% of intervention arm patients remained in care compared to 46% of standard arm patients. The timing of attrition from care was also consistent: many more standard arm patients (25% standard v 14% intervention) failed to initiate ART within 28 days, while somewhat more intervention arm patients were lost after initiation (35% intervention v 29% standard).

We traced viral load tests results between 9 and 14 months after enrolment for 365 (64%) of the 574 study patients retained in care through 14 months. Overall, viral load suppression was similar between the arms (30% versus 34% for standard and intervention arm patients, respectively). Viral suppression rates were high among those with a test result, with only 3% observed with an unsuppressed HIV viral load result in either arm. Table 1 presents two alternate outcome definitions for viral suppression: 1) observed viral load tests between 9 and 14 months after study enrolment to allow for the possibility that high rates of missing test results were due to the 12-month viral load test being done earlier; and 2) observed viral load tests done at any point up to 14 months after enrolment to measure differences in viral load suppression independent of ART initiation.

In this extended analysis of the SLATE I and SLATE II trial data from South Africa, we found that attrition from care at site by 14 months was high in both studies and both arms, but we found no evidence that the timing of ART initiation led to greater overall attrition one year after patients had the opportunity to start ART.

At the same time, it does appear that the offer of SDI shifted some attrition from before to after dispensing of the first dose of medication. For some intervention arm patients—those who were not in fact ready to start ART—it may have been easier to accept the offer of same-day initiation and drop out of care afterward than to refuse the offer when face-to-face with the care provider. We interpret this result to mean not that SDI “causes” post-initiation attrition from care, but rather that there is a certain proportion of patients who will drop out of care no matter how it is delivered, and an offer of same-day initiation will not change this fact[15]. What SDI can do, as we stated in an earlier publication, is “to prompt those who do make it to the clinic at least once to give ART a try, rather than being sent away empty-handed.”[16] Effective interventions to retain patients after they have started ART remain a high priority, regardless of the timing of initiation.

Recent observational studies of routine ART initiation have reported higher loss to follow up among patients who start ART on the same day as their HIV diagnosis than among those who do not.[17–19] While we cannot be certain of the reason for the discrepancy between our results and these studies’ findings, we speculate that there are two main explanations. First, it is likely that the patients offered and accepting SDI in routine service delivery differ from the ART-eligible patient population as a whole. For example, providers may offer SDI to patients whom they fear will not return for a second initiation visit, but these may also be patients who are at higher risk of post-initiation loss to follow up. Second, the SLATE algorithms included more than just the offer of SDI. Intervention arm patients participated in a structured preparation process implemented by trained study staff who may have been more successful than regular clinic providers in motivating patients to remain in care. The quality of the ART initiation process may thus be an important predictor of outcomes, along with the timing.

The SLATE trials had a number of limitations which are addressed in our previous reports [6,16]. The limitation most likely to have affected these results was our reliance on routinely collected data and the absence of unique identification numbers in the South African health system. This meant that we could not trace or ascertain the true outcomes who appeared to be lost to follow-up, some of whom likely remained on or re-started ART at other facilities. We have no reason to suspect that this limitation would have affected our study arms differentially, however.

## Conclusions

We conclude from this longer-term analysis of our SLATE I and SLATE II cohorts that an offer of same-day initiation of ART, following a carefully designed protocol to identify patients who are eligible and ready to start treatment, is not inherently associated with an overall increase in patient attrition from care.

## Data Availability

Data generated by the study will be posted in a public repository within one year of protocol closure. Data not owned by the authors, such as routinely collected medical record data, can be obtained from the owner, the South African National Department of Health.

## Supplemental digital content

Supplementary table 1. Results for SLATE II.

Supplementary table 2. Results for SLATE I.

## Authors’ contributions

MM, ATB, MPF, WDFV, and SR conceptualized the study and designed the protocol. LV, ATB, and MM collected and curated the data. MM, ATB, and MPF analyzed the data. MM and SR drafted the manuscript. All authors reviewed and approved the manuscript.

## Acknowledgments

We thank the patients who participated in the study and the staff of the study clinics for their cooperation and the City of Johannesburg and Ekhureleni Metro in South Africa. We also acknowledge the technical contributions of Dr Peter Ehrenkranz from the Bill & Melinda Gates Foundation.

