## Supplementary files for "Retention in care and viral suppression after same-day ART initiation: One-year outcomes of the SLATE I and II individually randomized clinical trials in South Africa"

**SUPPLEMENTARY MATERIALS**

**Supplementary table 1: Results for SLATE I**

| **Outcome** | **Standard arms (n=302)** | **Intervention arms (n=298)** | **Crude RD (95%CI)†** | **Crude RR (95% CI)†** |
| --- | --- | --- | --- | --- |
| **Previously reported outcomes** |  |  |  |  |
| Initiated ART ≤ 28 days of study enrolment | 204 (68%) | 232 (78%) | 10% (3 to 17%) | 1.15 (1.04 to 1.27) |
| Initiated ART ≤ 28 days and retained in care 8 months after study enrolment | 146 (48%) | 161 (54%) | 6% (-2% to 14%) | 1.12 (0.96 to 1.31) |
| Initiated ART ≤ 28 days and known to be virally suppressed by 8 months | 90 (30%) | 93 (31%) | 1% (-6% to 9%) | 1.05 (0.82 to 1.33) |
| **14-month outcomes (retention)*** |  |  |  |  |
| Initiated ART ≤ 28 days and retained in care 14 months after study enrolment | 153 (51%) | 149 (50%) | -1% (-9% to 7%) | 0.99 (0.84-1.16) |
| Initiated ART ≤ 28 days, not retained 14 months after study enrolment | 51 (17%) | 83 (28%) | 11% (4% to 18%) | 1.65 (1.21-2.25) |
| Did not initiate ≤ 28 days | 98 (32%) | 66 (22%) | -10% (-17 to -3%) | 0.68 (0.52-0.89) |
| **14-month outcomes (viral suppression)**** |  |  |  |  |
| Initiated ART ≤ 28 days and known to be retained and virally suppressed by 14 months | 65 (22%) | 68 (23%) | 1% (-5% to 8%) | 1.06 (0.79-1.43) |
| Initiated ART ≤ 28 days and known to be virally unsuppressed by 14 months | 8 (3%) | 2 (1%) | -2% (-4% to 0%) | 0.25 (0.05-1.18) |
| No viral load test results found | 80 (26%) | 79 (26%) | 0% (-7% to 7%) | 1.00 (0.77-1.31) |

*Observed clinic visit or VL test between months 11-14 after study enrolment

**Observed VL test between months 11-14 after study enrolment

**Supplementary table 2: Results for SLATE II**

| **Outcome** | **Standard arms (n=297)** | **Intervention arms (n=296)** | **Crude RD (95%CI)†** | **Crude RR (95% CI)†** |
| --- | --- | --- | --- | --- |
| **Previously reported outcomes** |  |  |  |  |
| Initiated ART ≤ 28 days of study enrolment | 243 (82%) | 277 (94%) | 12% (7% to17%) | 1.14 (1.08–1.22) |
| Initiated ART ≤ 28 days and retained in care 8 months after study enrolment | 175 (59%) | 220 (74%) | 15% (8% to 23%) | 1.26 (1.12–1.42) |
| Initiated ART ≤ 28 days and known to be virally suppressed by 8 months | 94 (32%) | 130 (44%) | 12% (5% to 20%) | 1.39 (1.12–1.71) |
| **14-month outcomes (retention)*** |  |  |  |  |
| Initiated ART ≤ 28 days and retained in care 14 months after study enrolment | 166 (56%) | 168 (57%) | 1% (-7% to 9%) | 1.02 (0.88-1.17) |
| Initiated ART ≤ 28 days, not retained 14 months after study enrolment | 77 (26%) | 109 (37%) | 11% (3% to 18%) | 1.42 (1.11-1.81) |
| Did not initiate ≤ 28 days | 54 (18%) | 19 (6%) | 12% (6% to 17%) | 0.35 (0.21-0.58) |
| **14-month outcomes (viral suppression)**** |  |  |  |  |
| Initiated ART ≤ 28 days and known to be retained and virally suppressed by 14 months | 111 (37%) | 115 (39%) | 2% (-6% to 9%) | 1.04 (0.85-1.28) |
| Initiated ART ≤ 28 days and known to be virally unsuppressed by 14 months | 11 (4%) | 13 (4%) | 0% (-2% to 4%) | 1.19 (0.54-2.60) |
| No viral load test results found | 44 (15%) | 40 (14%) | 1% (-7% to 4%) | 0.91 (0.61-1.36) |

*Observed clinic visit or VL test between months 11-14 after study enrolment

**Observed VL test between months 11-14 after study enrolment
